# Development of clinical virtual care pathways to engage and support families requiring neonatal intensive care in response to the COVID-19 pandemic (COVES Study)

**DOI:** 10.1101/2021.03.29.21254567

**Authors:** Marsha Campbell-Yeo, Justine Dol, Brianna Richardson, Holly McCulloch, Amos Hundert, Sarah Foye, Jon Dorling, Jehier Afifi, Tanya Bishop, Rebecca Earle, Annette Elliott Rose, Darlene Inglis, Theresa Kim, Carye Leighton, Sally Loring, Gail MacRae, Andrea Melanson, David C Simpson, Michael Smit, Leah Whitehead

**Affiliations:** School of Nursing, Dalhousie University, Halifax, Nova Scotia, Canada; Faculty of Health, Dalhousie University, Halifax, Nova Scotia, Canada; Department of Pediatrics, IWK Health & Dalhousie University, Halifax, Nova Scotia Canada; Centre for Pediatric Pain Research, IWK Health, Halifax, Nova Scotia, Canada; IWK Health, Halifax, Nova Scotia, Canada; Parent Partner, Neonatal Intensive Care Unit, IWK Health, Halifax, Nova Scotia, Canada; Nova Scotia Health Authority, Nova Scotia, Canada; School of Information Management, Dalhousie University, Halifax, Nova Scotia, Canada

**Keywords:** virtual pathways, co-design, COVID-19, neonatal intensive care

## Abstract

**Background:** In response to the COVID-19 pandemic, family presence restrictions in neonatal intensive care units (NICU) were enacted to limit disease transmission and protect infants, families, and healthcare providers. The effects of pandemic parental restrictions on providing optimal family integrated neonatal care is unknown.

**Aim:** To ensure optimal neonatal care using virtual care pathways to engage and support families in response to parental presence restrictions imposed during the COVID-19 pandemic. The research had two objectives: (1) conduct a needs assessment with families and healthcare providers (HCPs) of infants in the NICU to understand the impact of COVID-19 restrictions; and (2) develop virtual clinical care pathways to meet identified needs.

**Methods:** This study used focus groups and individual semi-structured interviews with families and HCPs for the needs assessment and identification of barriers and facilitators, and co-design for the development of the clinical virtual care pathways. For objective 1, content analysis was conducted by two independent reviewers to categorize findings and identify important barriers and facilitators of family-integrated care. For objective 2, an agile, co-design process utilizing expert consensus of a large interdisciplinary team was used to develop the care pathways.

**Results:** A total of 23 participants were included in the needs assessment (objective 1): 12 families and 11 HCPs. Themes identified were: (1) the need to maintain and build relationships and support systems; (2) challenges in accessing education and resources to integrate families in care; and (3) lack of standardized, accessible messaging related to COVID-19. For objective 2, we used the themes identified in the needs assessment to co-design three clinical virtual care pathways: (1) building and maintaining relationships between family and healthcare providers; (2) awareness of resources; and (3) standardized COVID-19 messaging.

**Conclusion:** Families reported that restrictive parental presence policies affected their mental health, well-being and social support. Families and HCPs reported the restrictions impacted delivery of family integrated care, education, transition to home, and standardized messaging. Clinical care virtual pathways were designed to meet these needs to ensure more equitable family centred care.

## 1 Introduction

In response to SARS-CoV-2 (COVID-19), public health restrictions were instituted worldwide to limit disease transmission and reduce burden on the health care resources and healthcare provider workforce.^1^ One such public health restriction was eliminating or severely restricting family or support person presence in hospital or clinic settings.^2,3^ Despite the known benefits of parental presence in the neonatal intensive care unit (NICU) settings, to protect the infants, families and healthcare providers (HCPs), most NICUs also instituted severe parental presence (visiting) restrictions.^2,3^

Each year, 15 million babies are born preterm, defined as less than 37 weeks gestational age, with the majority requiring neonatal intensive care.^4^ Worldwide and in Canada, prematurity is the leading cause of infant disability and death.^5^ While the youngest and sickest infants and their families are most at risk, even infants delivered one to two weeks early are at risk for immediate and long-term negative health outcomes. Most notably, infants are at risk of having developmental delay, and social, emotional and behavioural problems.^6,7^

Beyond infant outcomes, parents of these infants report higher levels of immediate stress, anxiety, depression, posttraumatic stress and greater adverse parenting outcomes than parents of healthy newborns.^8,9^ In order to improve the outcomes of vulnerable infants and their families and ease health care system burden, strong parental presence and education along with family integrated interventions have been shown to be a beneficial component of care in the NICU.^10–14^ Infant benefits associated with greater parental presence in the NICU include supportive effects for brain development,^15^ improved stress regulation,^16^ infant growth and a reduced length of stay.^17,18^ Greater parental presence and engagement in the NICU has also been associated with increased parental self-efficacy upon discharge,^19,20^ improved parent-infant relationships,^13,17^ decreased parental stress, depression, and anxiety,^11,12,21,22^ and increased exclusive breastmilk feeding at discharge.^10,23^

At IWK Heath, parental presence restrictions were instituted on March 13^th^, 2020 because of the local state of emergency declared due to COVID-19. IWK Health is a perinatal and pediatric university-affiliated referral hospital providing tertiary care to women and children across three provinces in Eastern Canada. The most severe restrictions limited only one support person to be present with their infant(s) who could not leave the hospital. If the women who delivered had a support person during delivery, the mother’s support person was required to leave 48 hours after delivery. These restrictions meant that families who stayed in the NICU with their infant(s) lacked access to their usual social support systems, partners had little to no access to their infant(s), and usual in-person teaching and access to resources were adversely affected. Additionally, some parents with responsibilities such as other children in the home had to leave the hospital, leaving their infant alone as the parent could not return to the NICU if they left.

Despite well intentioned public health decisions to limit the spread of COVID-19, many questions remain related to the impact of severe parental presence restrictions in the NICU on infant outcomes and parent mental health and well-being. Moreover, little is known regarding optimal ways to engage and support families in response to parental presence restrictions.

## 2. Study Team and setting

The Care Optimized using clinical Virtual pathways to Engage and Support NICU families in response to COVID-19 (COVES) study is comprised of a 20-member diverse interdisciplinary research team. The team includes parent partners (parents of infants requiring neonatal care), neonatal HCPs (neonatologists, neonatal nurse practitioners, nurses, educators, discharge planners, clinical nurse specialist), administrators (managers, directors and executive leaders) and researchers.

The study was conducted at IWK Health. The IWK Health NICU is a 40-bed single family room (with a designated sleep space, bathroom and shower) unit which provides level 3-4 care to 500-600 inborn and outborn patients annually. The NICU and hospital has a strong culture supporting family integrated care.

## 3 Methods

### 3.1 Objective 1: Needs Assessment

#### 3.1.1 Aim

The aim of objective 1 was to conduct a needs assessment with families and HCPs of infants in the NICU to understand the impact of the COVID-19 restrictions and determine care priorities to ensure optimal family-integrated care. Findings were anticipated to inform objective 2 of the study.

#### 3.1.2 Design

This study used a qualitative research design to conduct a needs assessment through focus groups and individual semi-structured interviews with families and HCPs. Ethical approval was received through IWK Health prior to recruitment (REB #1025748).

#### 3.1.3 Study population

English speaking families of an infant who was an inpatient in the NICU after March 13th, 2020 were considered eligible. For the purpose of the study, eligible family members were any primary caregiver of the infant. Families of infants who were palliative were not excluded from the study but were not actively approached.

Healthcare providers were considered eligible if they were providing clinical care in the IWK Health NICU since March 2020. There were no exclusions based on discipline or years of experience.

Purposeful sampling was also used to ensure a diverse sample, including underrepresented and equity seeking groups. We aimed to include a minimum of ten participants from each group (family and HCPs) or until data saturation was reached.

#### 3.1.4 Recruitment and Procedures

Virtual recruitment for the needs assessment occurred between May 21^st^, 2020 and June 29^th^, 2020. Given pandemic restrictions, usual in-person recruitment was not permitted. We recruited using posters in the NICU and through social media posts. Parents were also offered letters from the clinical team introducing the study and inviting them to participate. Interested families provided contact information and were contacted by a research coordinator by phone or email. Healthcare providers were recruited through letters of invitations and interested HCPs provided their contact information agreeing to be contacted by the research coordinator. All participants provided written or e-consent and completed an online demographic form. Focus groups and interviews were conducted via the Zoom video conferencing platform. Participants were provided a secure password protected Zoom link to join a focus group or interview (based on participant choice). Sessions were between 40 and 60 minutes in length and were recorded and transcribed. Participants were informed of ways to receive support should the content of the focus group discussion or interview caused them stress or anxiety.

The semi-structured interview guide (supplementary material) was created through existing evidence and expert consensus of the full research team, comprised of parent partners from eastern Canada and neonatal clinicians, administrators and researchers who work at the IWK Health NICU.

#### 3.1.5 Analysis

The interviews and focus groups were analyzed using inductive thematic qualitative content analysis.^24^ Content analysis was conducted using NVivo 12 by two independent reviewers to categorize findings and identify the impact of restrictive parental presence polices in the NICU in response to COVID-19. A third reviewer resolved any disagreements and assisted with creation of the themes. Follow-up and participant feedback were utilized to ensure credibility and trustworthiness of the data.^25^

To determine potential barriers and facilitators for development and practice uptake, interviews and focus groups were analyzed using qualitative content analysis and the domains of the Theoretical Domains Framework (TDF)^26^ by two reviewers to categorize findings and identify important barriers and facilitators. Barriers and facilitators to implementing virtual care pathways during COVID-19 were identified using the Behaviour Change Wheel (BCW)^27^ and TDF.^26^ Priorities for pathway creation were identified with final suggestions for revisions reviewed by the research team and changes made based on consensus.

### 3.2 Results

A total of 23 participants (12 family and 11 HCP) participated. All participants were included in the analysis. Nine interviews were conducted with family participants and data from the HCPs were collected through two focus groups and one interview. Family participants included 9 mothers, 2 fathers and 1 grandfather. Healthcare provider participants included 3 registered nurses, 1 neonatologist, 2 neonatal nurse practitioners, 2 lactation consultant specialists, and 2 pharmacists. All but 2 of the HCPs were female.

Overall, while families reported experiencing significant distress, all reported feeling confident in the quality of care their infant received. Identified needs focused almost exclusively on parent outcomes related to emotional and physical well-being. Three primary areas emerged that guided the development of the pathways: (1) the need to maintain and build relationships and support systems; (2) challenges in accessing education and resources to integrate families in care; (3) lack of standardized and accessible messaging related to COVID-19 pandemic response.

#### The need to maintain and build relationships and support systems

Parental presence restrictive policies adversely impacted parent and HCPs therapeutic relationship, parent-family relationships and access to usual social support systems. Parent mental well-being was negatively impacted by the restrictions of limiting the number of primary support persons, not allowing primary support persons to come and go, and preventing siblings and extended family members from visiting. Families reported feeling disconnected from their infant and the care team. Healthcare providers similarly reported feeling disconnected from families as well as other HCPs. Concerns were raised regarding the ability to establish therapeutic relationships with families.

#### Challenges in accessing education and resources to integrate families in care

Healthcare providers reported that their ability to maintain and foster family integrated care was adversely affected. Both parents and HCPs reported concerns related to families accessing educational materials, in-person HCP teaching, and increased burden on some family to teach other family members at home. This concern was particularly relevant regarding discharge planning.

Healthcare providers reported concerns with both themselves and families being left out of usual care decisions made during daily medical rounds when the number of people permitted at the patient bedside was reduced as part of the physical distancing policies. Families and HCPs also reported challenges pivoting to virtual care. Attempts to include HCPs and families using telephone or video conferencing was associated with concerns related to poor audio quality, fear of errors being made and issues related to privacy. Issues related to equitable care were raised, primarily regarding family access to reliable Wi-Fi or devices, and comfort with the use of virtual resources. Learning needs regarding comfort with technology during rapid pivot to virtual platforms were also reported.

#### Lack of standardized and accessible messaging related to COVID-19 pandemic response

Significant need for standardized, easily accessible COVID-19 messaging in the NICU and across the women’s newborn program for both families and care providers were identified. Unmet parental physical needs were also reported which included difficulties acquiring necessary prescriptions, personal items, and clothing. These concerns were greatest in non-local residents (where closed borders between provinces precluded travel), and immigrant populations.

### 3.2 Objective 2: Clinical virtual care pathways

#### 3.2.1 Aim

The aim of objective 2 was to develop clinical virtual care pathways to meet identified needs from objective 1.

#### 3.2.2 Study Design

Agile, collaborative co-design process using expert consensus of interdisciplinary team which included parents, and qualitative interviews with families, clinicians, and decision-makers.^28^

#### 3.2.3 Study Population

A diverse 20-member interdisciplinary research team which included parents of previous NICU patients participated to provide expert consensus recommendations on the development of the co-designed pathways. Qualitative interviews and iterative testing were conducted with families and HCPs to further adapt and revise pathways. Eligibility criteria were consistent with objective 1. Eligible participants were the intended users of the clinical virtual care pathways, HCPs from the NICU of IWK Health in Eastern Canada with inclusion and exclusion criteria as noted above.

#### 3.2.4 Recruitment and Procedures

Recruitment and co-design sessions for objective 2 occurred between July 14^th^, 2020 and December 1^st^, 2020. To aid in the development of the pathways, weekly smaller working groups and full team bi-monthly 60-90 minutes virtual interdisciplinary research team meetings were held. For the purpose of identification of barriers and facilitators and iterative co-design testing cycles, we used the same recruitment procedures utilized in objective 1. Participants were prompted using a semi structured interview guide (supplementary material). Iterative bi-monthly agile co-design sessions and live online documents available to the full research team were utilized to further adapt and revise the co-designed pathways. Additional need for iterative agile sessions were determined based on saturation of data obtained.

#### 3.2.5 Analysis

Interviews and focus groups were analyzed using qualitative content analysis and the domains of the TDF by two reviewers to categorize findings and identify important barriers and facilitators. Barriers and facilitators to implementing virtual care pathways during COVID-19 were identified using the BCW and TDF.^26,27^ All interviews were conducted using a semi-structured interview guide based on the TDF, created by the full research team. Domains were mapped on to virtual care functions to guide options for changes. The APEASE intervention criteria of the BCW (affordability, practicability, effectiveness and cost-effectiveness, acceptability, safety, and equity) informed decision making.^27^ Priorities for pathway creation were identified, and recommendations for revisions were reviewed by the research team and changes were made based on consensus.

### 3.3 Results

#### 3.3.1 Pathway Overview

The pathways document a step-by-step process to ensure standardized care is offered to families, addressing the themes identified in objective 1. While the primary users are likely to be families and bedside nurses, the entire multidisciplinary care team (i.e., pharmacists, lactation consultants, advanced practice nurses and physicians) are encouraged to complete the relevant section of the pathways. The pathways incorporate an algorithmic approach that includes decision points to take into consideration family scenario and timing. The main introduction page for the pathways (Figure 1) includes a description of the potential family scenarios that should be considered for each element of the pathways. This step aims to ensure that all identified primary parent/caregivers would be contacted regardless of their ability to be present in the NICU. All elements of the pathways are allocated to one of the following time frames to ensure parent’s received timely information: (1) admission; (2) during stay; or (3) prior to discharge. HCP documentation associated with each step of the pathways allows HCPs to keep track if the pathway element was offered regardless of location of family and whether the step was utilized by families. Follow up prompts are embedded in the pathways to determine if the families have questions. Educational resources are offered in a virtual format accessible at any time from hospital or from home. The pathways provide a standardized approach to directing parents to virtual resources as part of usual care. Virtual resources utilized are part of an existing web-based evidence based educational platform called Chez NICU Home (CNH), recently developed at IWK Health, planned to be fully implemented in the NICU prior to the COVID-19 pandemic. Pandemic research restrictions altered the planned research implementation and practice uptake of the resource. The platform includes interactive parent training, and education. The CNH platform was developed following user-centered principles of design including iterative testing and feedback from parent and HCP end users. The platform consists of six core chapters which include lessons and resource pages broken down into reading resources and video content. There is also a 12-step Discharge Planning Module, Progress Dashboard, and Interactive Tracker to record parental presence and involvement in care. The platform includes 14 instructional videos teaching key parental skills such as car seat safety and pumping breastmilk. The virtual communication platform utilized in the pathways was created to align with the CNH platform and was referred to as the CNH Connect. The CNH Connect utilizes a secure WebX video/audio platform that can be accessed via multiple devices (phone, tablet, in room screen, or computer) from both in and outside the hospital. The CNH Connect was not available to families and staff prior to the pandemic restrictions.

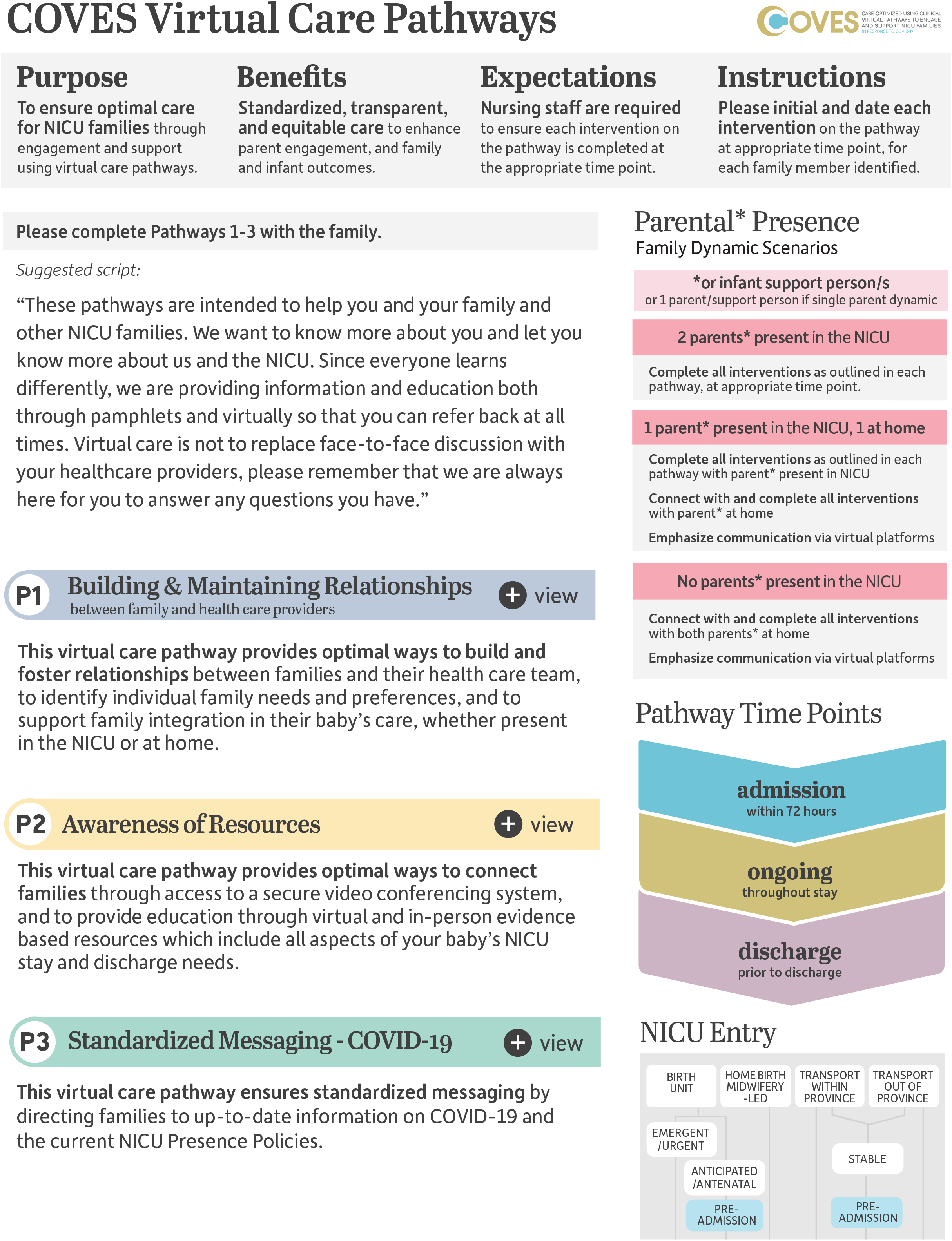

#### 3.3.2 Clinical Virtual Care Pathways

The three clinical virtual care pathways align with the themes identified in objective 1: (1) building and maintaining relationships between family and HCPs; (2) awareness of resources; and (3) standardized messaging related to COVID-19.

##### Pathway #1 Building and maintaining Relationships

The aim of the pathway is to get to know the family. In doing this, we can provide optimal virtual ways to build and foster relationships between families and their health care team. This helps to identify individual family needs and preferences, and to support family integration in their baby’s care, whether present in the NICU or at home (Figure 2).

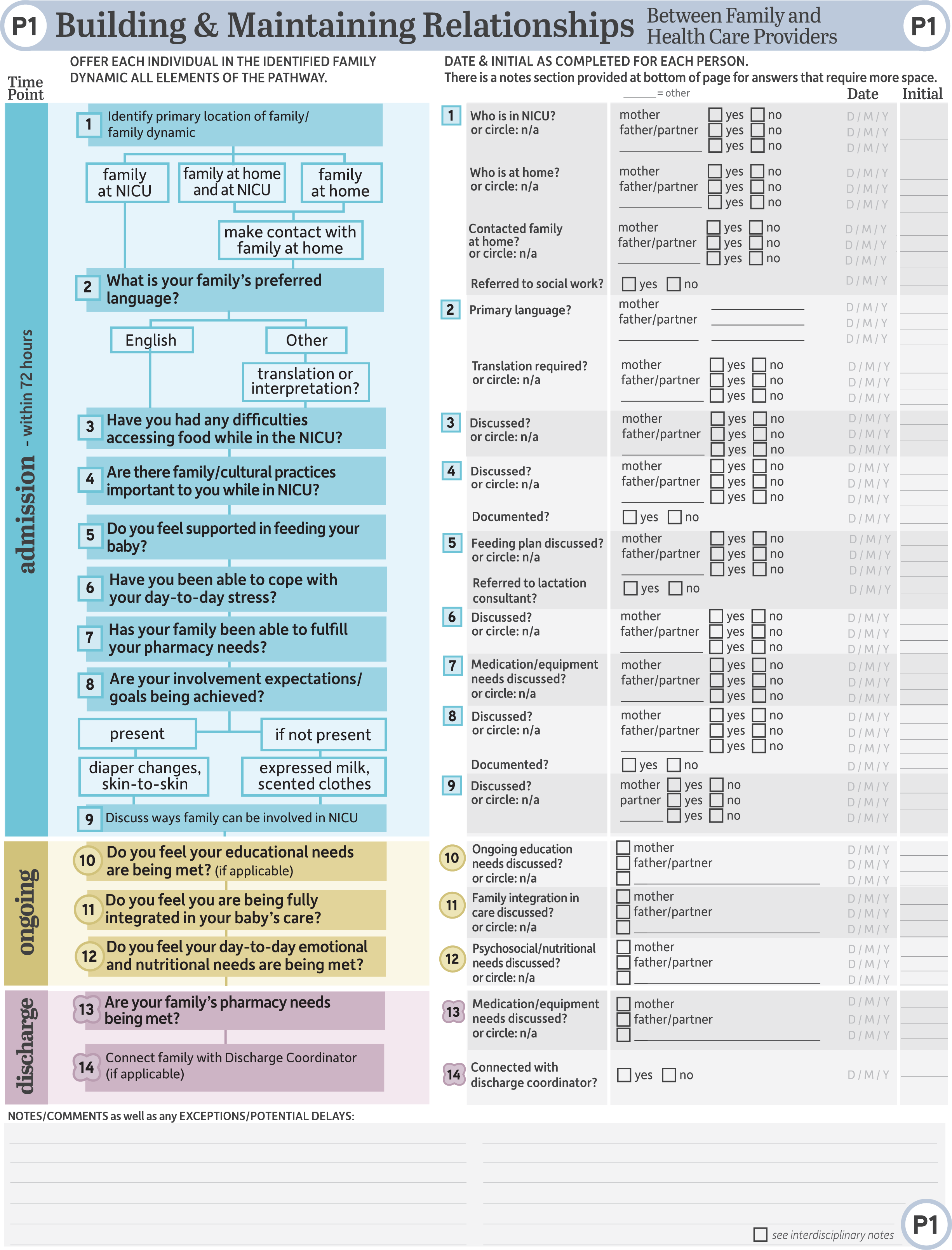

##### Pathway #2 Awareness of Resources

The aim of this pathway is to ensure that families have equitable awareness and access to evidence-based resources both virtually and in person. The pathway provides optimal ways to connect families through access to a secure video conferencing system, and to education through virtual and in-person resources which include all aspects of the infant’s NICU stay and discharge needs (Figure 3).

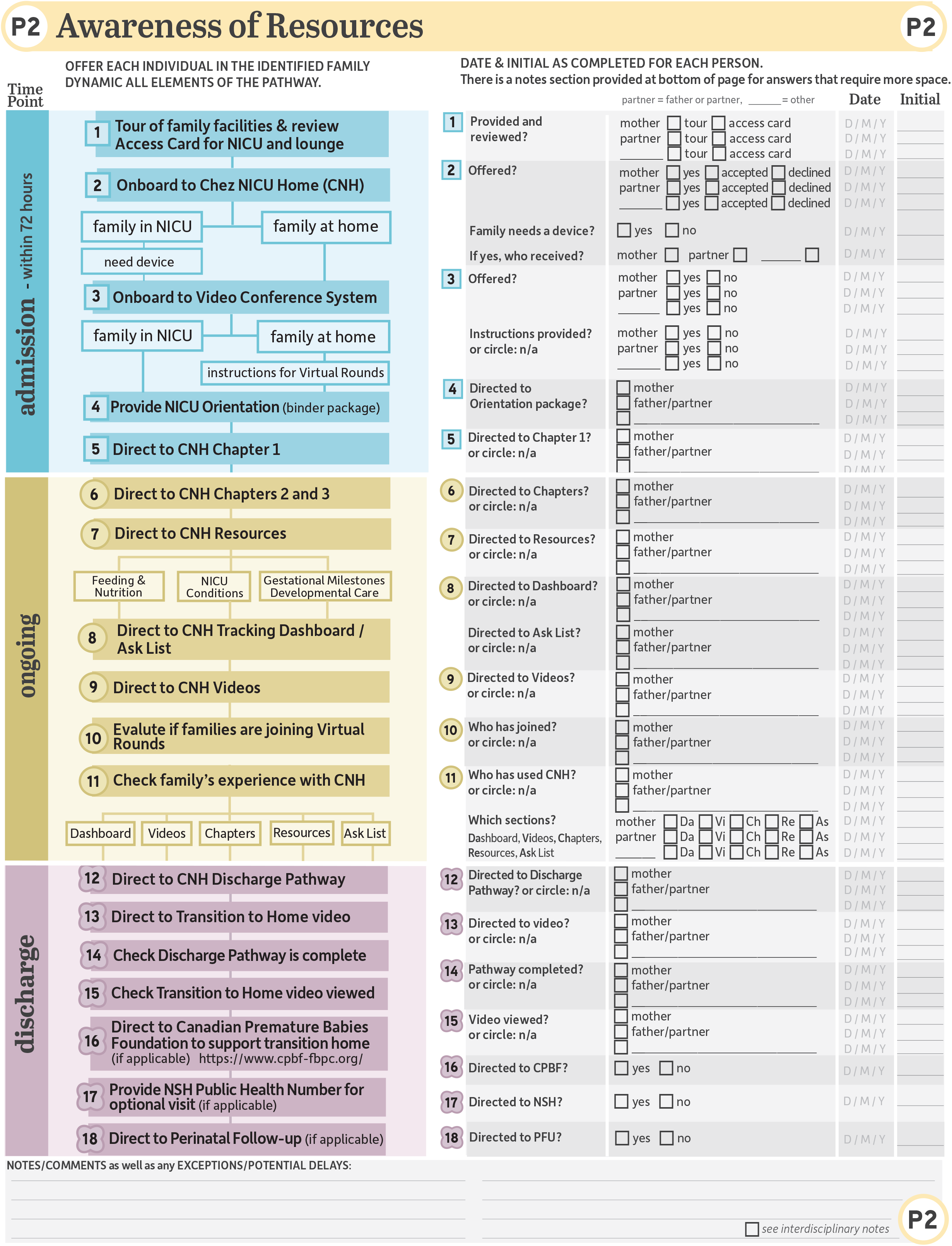

##### Pathway #3 Standardized Messaging – COVID-19

The aim of this pathway is to address the rapid changes to public health guidelines and institutional policies as a result of COVID-19. This will ensure families and HCPs have better and consistent access to the standardized messaging they need to navigate through the NICU presence policies and receive the most current information on COVID-19 (Figure 4).

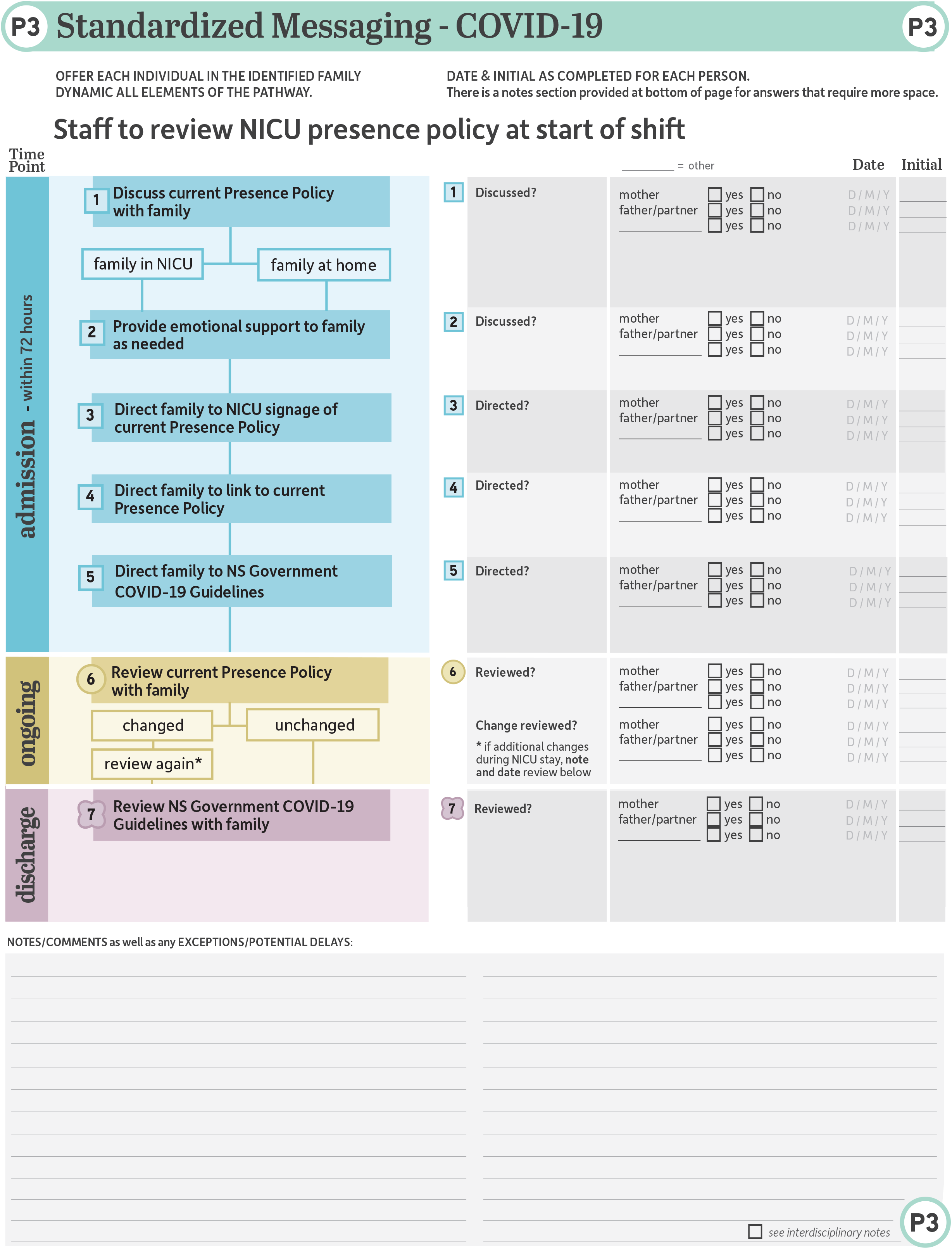

##### Barriers and facilitators to practice uptake

To help determine barriers and facilitators to pathway acceptance and feasibility for implementation and practice uptake, we interviewed ten participants between October 25^th^, 2020 and November 25^th^, 2020. This included one family member (mother) and nine HCPs (7 nurses; 1 neonatal nurse practitioner; and 1 pharmacist).

The participants reported feeling somewhat overwhelmed when initially seeing the pathways but when provided opportunity to review the pathways, they agreed that all of the elements were relevant. There were some suggestions for wording changes related to equity and offering devices to families in NICU to use for resources and attendance at rounds. Wording changes to put more emphasis on the purpose for virtual care and transparency, clarity around exceptions and an emphasis on ensuring families have access to food were also suggested.

Most HCPs felt confident to complete the steps of the pathway. Some identified that aspects of the pathways required additional information and resources, most notably standardized messaging to families and additional training related to onboarding families to virtual communication and educational resources. Concerns were raised around the potential increased nursing workload. However, participants identified family questions as part of the existing charting admission process that could be potentially integrated as part of the pathways to streamline care and reduce duplication.

Families felt that pathway elements were relevant and time frames were reasonable. They identified the importance of a space to record reasons for delay or exceptions to completing any elements of the pathways. Families also reported the importance of ensuring that HCPs did not pressure families by prescribing the pathways but direct the families to the resources, allowing them to freely make the decision about whether to use them or not. Families raised the importance of completing the pathways “with families” rather than “to families,” as well as the need to be more inclusive of fathers.

##### Iterative Pathways revisions

Changes to the pathways were made based on user responses and expert consensus of the full co-design research team input and co-design sessions. The following changes or additions were made to inform final pathways.

To inform learning needs and standardized messaging, a user guide (see supplemental materials) and a script (main page of pathways (Figure 2) were created and added. To emphasize the integration of families, elements of the pathways were rephrased as questions using simplified language versus using statements. The terminology “father /partner” was added versus only partner to be more inclusive of fathers.

To reduce nursing workload, existing family targeted questions from the unit’s current admission documentation was removed and incorporated as part of the pathways. Information was added to clarify that all care providers should participate in completing relevant aspects of the pathways. For example, the lactation consultant directing a family to the online education module and video regarding pumping breastmilk and safe storage, or the pharmacist directing a family to view the online video about giving medications. It was decided that unit ward clerks would assist with the on-boarding process for the educational (CNH) and communication (CNH Connect) platforms. All staff will be provided training on use of the pathways in a 14 day in-person roll-out and would be provided access to written and online resources (user guide and training presentation).

### 3.4 Discussion

We aimed to determine the impact of parental presence restrictions in response to the COVID-19 pandemic on parents of infants requiring neonatal intensive care, as well as develop clinical virtual pathways to address identified needs of families to better engage and support families to be fully integrated as part of their infant’s care team. The development of the virtual clinical care pathways was done through an agile, collaborative process based on the needs identified from NICU families and HCPs. To enact meaningful change, decisions on the format of these pathways were responsive to the needs of families and pragmatic in considering current unit practices. As such, the pathways were formatted to use a low-tech approach of an interactive PDF, while the contents of the pathways standardized the use of virtual solutions for optimizing clinical care.

#### 3.4.1 Needs Assessment

Our findings indicate the substantial impact that families experienced during severe parental presence restrictions instituted in response to the COVID-19 pandemic, consistent with a recent global survey of 277 facilities that reported a significant reduction on parental presence and engagement across NICUs.^2^

Interestingly, few families and HCPs expressed concerns regarding the impact of COVID-19 parental presence restrictions on infant outcomes, while all emphasized the impact on parent/family well-being. Identified needs focused almost exclusively on parent outcomes related to emotional and physical well-being. Our findings are in keeping with previous reports of altered family well-being during an infant’s NICU stay irrespective of COVID-19,^11,12,21,22^ as well as heightened concerns during the pandemic.^29^ Nevertheless, this is somewhat surprising given the known benefit of parental presence on infant outcomes.^15–18^ This may reflect a significant level of confidence in the care provided by the NICU team, a lack of insight or underappreciation regarding the important contribution of parental presence on infant outcomes, or simply that the degree of personal parent/family distress was too great to move beyond. Further inquiry is warranted to elucidate this finding.

#### 3.4.2 Co-Design of Clinical Care Pathways

The pathways were co-designed with parents, families, clinicians, decision-makers, and researchers based on findings from the needs assessment and continuously informed by the COVES team. Collaboration and creative feedback/input from the large, diverse COVES team and NICU families/current NICU staff/HCPs were essential components to the design stage to create a resource that would be relevant to practice and have a high likelihood of acceptance and uptake when incorporated into clinical practice to optimize care.

Co-design and participatory design have been demonstrated as an effective method for development of virtual and eHealth resources, specifically in the perinatal population,^30^ and in designing clinical care pathways to meet the unique needs of specific populations.^31^ The co-design/participatory design approach aligns with the philosophy of family-integrated care (upheld by the study institution) in that the health system is adapted to the specific needs of its population, and the only true way to achieve that is with the input of the population.^29–32^

#### 3.4.3 Pivoting to Virtual Care

Pivoting to virtual care was necessary to optimize clinical care during the period of uncertainty and in-person interaction limitations due to parental presence restrictions enforced for the pandemic. The value of virtual care was quickly acknowledged by many in healthcare systems and widely pursued to maintain support and care for the community.^33–35^

In response to the needs assessment and need for a development of an intervention that was tailored to our institution/NICU, we decided to first create the suite of pathways in paper form and as interactive PDF to ensure these pathways could easily be adopted into clinical practice and patient documentation. The initial paper version allowed for rapid and responsive revisions to be made during development as well as throughout the initial roll out and implementation. The pathways currently will exist as part of the paper-based charting system but will also be housed digitally as the interactive PDF for information purposes.

Throughout this study, the process of designing the clinical care pathways acted as a catalyst for greater uptake of existing virtual care platforms within the NICU and across the institution. As previously mentioned, IWK Health has already integrated prior to the pandemic CNH and CNH Connect within their NICU, which is a virtual, eHealth education and connection platform. Thus, the virtual pathways offer the opportunity to systematize the introduction of the CHN platform to incoming patients, particularly in pathway 2. For example, in this pathway, there is increased opportunity for uptake of CNH through standardized onboarding practices with families in the NICU or at home. Additionally, it triggered rapid implementation of the CNH Connect to ensure family at home were incorporated into their infant’s care and decision making through virtual rounds. In the beginning of the pandemic and presence restrictions, families joined rounds virtually through technologies like FaceTime and Zoom, yet due to the critical nature of this environment and need for high quality infection control practices, any equipment (e.g., computer for Zoom) could not enter patient rooms. This barrier was shared across adult ICUs, which became a greater issue as many hospitals restricted family presence entirely.^36^ Expediating the rollout of CHN Connect enhanced virtual rounds significantly, as it allowed families and other HCPs to easily join virtual rounds, enhanced patient privacy, and enhanced care through more effective communication. Other critical care areas utilized similar systems/platforms to facilitate presence when physical presence of families was restricted, which was reported to be incredibly impactful to the whole family unit specifically with their mental wellbeing.^36^

#### 3.4.4 Strengths and Limitations

The primary strength of this project was that the pathways were designed using user-centered design strategies.^37^ This included the needs assessment to define user/target audience, co-creation sessions (co-design), designed occurred through collaboration with multiple teams, the use of focus groups to gain user perspective, and iterative developments. As the goal of this project was to create a solution to optimize clinical care during the pandemic, it was imperative that we designed evidence-based care pathways. Dopp and colleagues state that applying a variety of user-centered design strategies facilitates the development of “evidence-based practice services, technologies, and implementation plans that address identified needs” (p.1059).^37^

However, despite this strength, there are some limitations that must be acknowledged. First, due to the constantly changing nature of the guidelines related to COVID-19 policies, difficulties emerged in connecting and engaging with users throughout this process. Difficulties arose recruiting families to participate as family members had to initiate contact with the study team to begin the virtual recruitment process. Another challenge was the implementation of the pathways as a completely virtual tool. Due to the iterative nature needed in the design and development, the pathways were initially piloted as a paper copy with transition occurring into an interactive PDF document for the ongoing pilot testing. Future incorporation into the CNH platform is desired.

## 4 Conclusion

COVID-19 policies restricted family presence and involvement in the NICU. Families reported that this affected their mental health, well-being and social support. Both families and HCPs reported the restrictions significantly impacted usual in-person parent integration in care, parent education and transition to home. Rapid change and variation of restrictive presence policies across multiple women and newborn departments and lack of easily accessible standardized messaging regarding polices were identified by both families and HCPs as a highly stressful and significant concern. Virtual clinical care pathways were developed to meet these needs and preliminary feedback suggests that the clinical implementation of these pathways will ensure more equitable family centred care across all parent scenarios regardless of whether they are in the hospital and/or at home.

## Data Availability

The data that support the findings of this study are available from the corresponding author, [MCY], upon reasonable request.

## 6 Author Contribution

Author MCY conceptualized the manuscript and wrote the first draft of the manuscript and edited all revisions. HM, BR, JD, AH and SF contributed substantial content to the paper. All co-authors were actively involved in all aspects of the study and creation of the pathways. All co-authors approved the paper.

## 8 Contribution to the field

Irrespective of a pandemic, the majority of parents of preterm infants experience high levels of stress, uncertainty, challenges navigating their role in the NICU and ability to be fully integrated as part of their infant’s care. Our study demonstrated the significant heightened impact of COVID-19 parental presence restrictions on parent well-being and identified gaps in care and identified key priority areas of need to promote family integrated care practices and engagement during the pandemic. The creation of clinical virtual pathways provided a standard and equitable approach to provide virtual family integrated care, including parental education and training while COVID-19 restrictions have limited our ability to deliver care face-to-face. Our findings will inform the feasibility, acceptably, and impact of prescribing a virtual care platform to deliver education and support to families of infants admitted an NICU within a family integrated care model. Evaluating the implementation and effectiveness of the pathways into care is warranted. In the future, beyond the COVID-19 pandemic, continued use of the care pathways may also have long term benefits particularly for rural, remote, and vulnerable families who are unable to ensure ongoing presence in the NICU.

